# Prediction of incident coronary artery disease in individuals with zero coronary artery calcium using a novel multi-ancestry, label-free polygenic risk score framework

**DOI:** 10.64898/2026.03.02.26347474

**Authors:** Giordano Bottà, Muriel Rossi, Jen Kintzle, Paolo Di Domenico

## Abstract

**Background:** A coronary artery calcium (CAC) score of 0 is widely considered to indicate low short- to intermediate-term risk for coronary artery disease (CAD) and is frequently used to defer lipid-lowering therapy. However, a subset of individuals with CAC=0 still experience events, highlighting residual risk not captured by imaging alone. Polygenic risk scores (PRS) quantify lifelong inherited susceptibility, but conventional approaches rely on predefined ancestry labels despite human genetic diversity existing along a continuum. To address this limitation, we developed *8 Billion*, a novel, label-free framework that models genetic similarity without pre-labeling individuals by ancestry. We evaluated whether a CAD PRS derived using this approach identifies clinically meaningful residual risk among individuals with baseline CAC=0.

**Methods:** We analyzed participants from the Multi-Ethnic Study of Atherosclerosis (MESA) with baseline CAC=0. The 8 Billion framework estimates individualized PRS by anchoring each participant to a genetically similar reference neighborhood rather than discrete ancestry groups. Multivariable Cox proportional hazards models assessed associations between PRS-defined risk groups and incident CAD, adjusting for principal components of genetic variation (PC1–PC4), age, sex, smoking status, systolic blood pressure, total and high-density lipoprotein cholesterol, diabetes, and antihypertensive medication use. Two classifications were evaluated: (1) a Top 5% group defined by the highest 5% of PRS-derived odds ratios in the cohort; and (2) an individualized high-risk group defined using a personalized threshold derived from the 8 Billion framework. Ten-year absolute risk estimates were derived from adjusted models.

**Results:** Despite CAC=0 at baseline, polygenic burden was independently associated with incident CAD. Individuals in the Top 5% PRS group had higher risk of CAD events compared with the remainder (hazard ratio [HR], 3.12; 95% CI, 1.05–9.31; P=0.041). The individualized high-risk group defined through 8 Billion was similarly associated with increased CAD risk (HR, 2.52; 95% CI, 1.12–5.66; P=0.025). Estimated 10-year ASCVD risk among high-PRS individuals exceeded the 7.5% threshold commonly used to guide initiation of lipid-lowering therapy, despite CAC=0. In fully adjusted models, conventional risk factors were not statistically significant within this subset.

**Conclusions:** Among individuals with CAC=0 in a multi-ethnic cohort, a label-free, ancestry-continuum PRS approach identified subgroups at significantly increased risk of incident CAD and at guideline-relevant 10-year treatment thresholds. Integration of polygenic risk with CAC imaging refines preventive decision-making beyond imaging alone.

**Clinical Perspective:** *What is new?:*

- Among individuals with baseline CAC=0, the Allelica Multi-ancestry CAD PRS calculated with the 8 Billion framework identified subgroups at significantly increased risk of incident CAD.
- In this CAC=0 population, high polygenic risk was associated with 10-year risk estimates above the 7.5% treatment threshold, whereas conventional risk factors were not statistically significant in adjusted models.

*What are the clinical implications?:*

- A CAC score of 0 should not be interpreted as uniformly protective, because genetically high-risk individuals may still experience clinically meaningful coronary events.
- Integrating PRS with CAC assessment may improve preventive decision-making by identifying patients with residual risk despite reassuring baseline imaging.
- In selected patients with CAC=0, high polygenic risk may support closer follow-up and earlier consideration of lipid-lowering therapy or other preventive strategies and imaging modalities.

## Introduction

Coronary vascular disease (CVD) remains the leading cause of death worldwide [1], with coronary artery disease (CAD) representing its most common manifestation. CAD reflects the combined effects of inherited susceptibility and environmental exposures [2,3]. Contemporary primary prevention guidelines recommend multivariable risk estimation using tools such as SCORE2 [4], the Framingham Risk Score [5], and the Pooled Cohort Equations (PCE) [6] to guide initiation of lipid-lowering therapy. However, a substantial proportion of individuals who ultimately experience a first coronary event are not identified as high risk by these conventional algorithms [7]. Coronary artery calcium (CAC) scoring, introduced by Agatston and colleagues [8], provides a noninvasive measure of calcified plaque burden using noncontrast computed tomography. Its prognostic value has been consistently validated across prospective cohorts, with strong and independent associations with coronary heart disease, stroke, and mortality [9, 10, 11, 12]. Current prevention guidelines recommend CAC scoring to refine statin allocation in individuals at borderline or intermediate estimated risk. A CAC score of 0 is widely regarded as a negative risk marker and is frequently used to defer statin therapy, with reassessment after 5 to 10 years [13, 14, 15, 16]. Nevertheless, clinical events continue to occur among individuals with CAC=0, underscoring residual risk not captured by calcium imaging alone [17]. Polygenic risk scores (PRS) quantify lifelong inherited susceptibility to CAD and represent a potential risk stratifier within individuals considered low risk by CAC imaging. By capturing cumulative genetic burden present from birth, PRS may identify individuals with elevated underlying risk despite absence of calcified plaque at a single time point. However, most existing PRS methods rely on predefined ancestry categories. This approach assumes discrete genetic groupings, whereas human genetic diversity exists along a continuum shaped by admixture and migration [18]. Categorizing individuals into rigid ancestry labels may therefore obscure genetic similarity and reduce performance in admixed populations. To address this limitation, we developed 8 Billion, a label-free framework that models genetic similarity as a continuum and derives individualized PRS estimates without assigning individuals to fixed ancestry groups. We applied this approach in the Multi-Ethnic Study of Atherosclerosis (MESA) [19], a prospective cohort specifically designed to include diverse and admixed populations and characterized by serial CAC measurements over time. Using this dataset, we investigated whether the Allelica multi-ancestry CAD PRS, estimated with 8 Billion, identifies residual coronary risk among individuals with CAC=0 at enrollment and refines risk stratification beyond imaging-defined low risk [20].

## Methods

The analysis was conducted in approximately 5,800 participants from the Multi-Ethnic Study of Atherosclerosis (MESA). After application of the following inclusion and exclusion criteria, the final analytic sample comprised 1,700 individuals: participants were required to have a baseline CAC score of 0 at study entry (T0) and at least 1 follow-up CAC measurement to permit longitudinal assessment. Individuals classified as East Asian or Admixed American were excluded because, after filtering, event counts were insufficient for reliable estimation. The final cohort therefore included 1,700 participants of African, European, or admixed ancestry with CAC=0 at enrollment and ≥1 follow-up scan. To obtain individualized and calibrated risk estimates, we developed and applied the 8 Billion framework. For each MESA participant, a genetically similar reference neighborhood was identified within the UK Biobank dataset. Genetic similarity was quantified using a kinship-based distance matrix, ranking reference individuals by ascending genetic distance from the target participant. Neighborhood size was optimized through incremental expansion. At each candidate size, a logistic regression model was fitted using PRS as the primary predictor, with adjustment for ancestry and clinical covariates. Model performance was assessed using the Brier score and stabilized via 1,000 bootstrap resamples with 90% subsampling. Validation subsets in which disease prevalence deviated by >1 SD from ancestry-specific means were excluded. Candidate neighborhoods were constrained to include ≥500 individuals. The final neighborhood size minimized the Brier score, optimizing calibration and discrimination for each individual. In parallel, a conventional continental-ancestry PRS model was computed for comparison. Using calibrated PRS estimates, analyses were restricted to participants with CAC=0 at baseline to evaluate the independent contribution of genetic risk in an imaging-defined low-risk population. Two PRS-based stratifications were prespecified: (1) Top 5%, defined as individuals whose PRS-derived odds ratios ranked in the highest 5% of the cohort; and (2) high-risk PRS, defined by an individualized ≥2-fold risk threshold. Ten-year CAD risk was estimated using Cox proportional hazards models adjusted for Pooled Cohort Equation variables. Kaplan–Meier analyses were performed to compare event-free survival across PRS strata over a mean follow-up of 10 years. Survival estimates reflect incident events after enrollment among participants free of clinical cardiovascular disease at baseline. A secondary analysis evaluated CAC progression to ≥100 using the same PRS stratifications to assess whether elevated genetic risk was associated with subsequent subclinical calcification despite baseline CAC=0.

## Results

Among participants with CAC=0 at baseline, individuals in the highest PRS strata demonstrated substantially greater estimated 10-year risk of CAD despite identical imaging-defined risk. Using the 8 Billion framework, both stratification approaches identified groups whose predicted 10-year risk exceeded the 7.5% threshold commonly used to guide lipid-lowering therapy [21], whereas individuals with lower PRS remained below this benchmark (Figure 1). Estimated 10-year CAD risk was approximately 9% in the Top 5% PRS group and 8% in the broader high-risk PRS group, compared with ≈4% among the remainder. Across the entire cohort without PRS stratification, mean 10-year risk was 4.7%, substantially lower than that observed in the highest PRS categories. Kaplan–Meier demonstrated reduced CAD event–free survival among genetically high-risk groups over the full study follow-up. Event-free probability declined to ≈0.85 in the Top 5% PRS group versus ≈0.94 in the remainder (log-rank P=0.03) and to ≈0.88 versus ≈0.94 in the high-risk PRS group (P=0.010) (Figure 2). These findings indicate sustained divergence in cumulative event rates over time among individuals classified as genetically high risk despite absence of baseline calcification. In multivariable Cox models adjusted for principal components (PC1–PC4), sex, smoking, systolic blood pressure, total and HDL cholesterol, diabetes, and antihypertensive therapy, CAD PRS remained independently associated with incident CAD. Individuals in the Top 5% PRS group had a threefold higher hazard of CAD compared with the remainder (hazard ratio [HR], 3.12; 95% CI, 1.05–9.31; P=0.041). Using the individualized high-risk PRS definition yielded consistent results (HR, 2.52; 95% CI, 1.12–5.66; P=0.025). Conventional risk factors were not statistically significant in these models, underscoring that PRS captured risk not explained by standard covariates within this CAC=0 subset.

**Figure 1.**
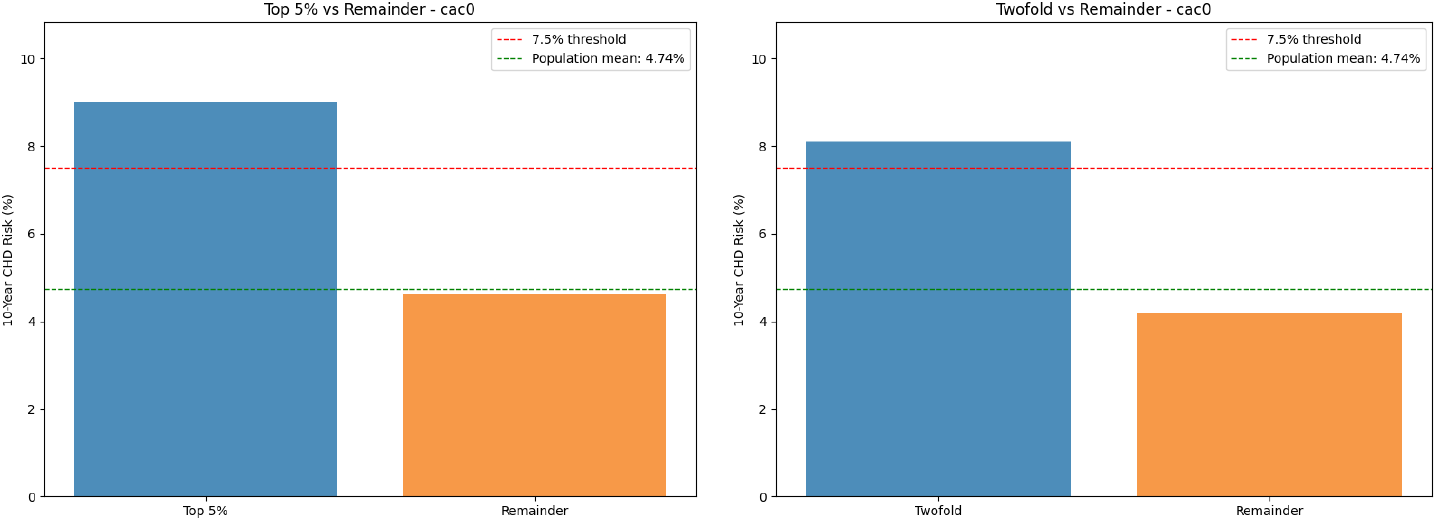
Ten-year coronary artery disease (CAD) risk stratification in a multi-ancestry cohort. Bar graphs display adjusted 10-year CAD risk estimates derived from Cox proportional hazards models including traditional cardiovascular risk factors and principal components of ancestry. Analyses were restricted to participants aged ≥45 years with baseline CAC=0 and available longitudinal follow-up. Left panel: Participants in the Top 5% PRS group had an estimated 10-year CAD risk of 9.02% compared with 4.62% in the remainder of the cohort. Right panel: Participants classified as high risk (PRS ≥ individualized threshold) had an estimated 10-year CAD risk of 8.10% compared with 4.19% in the remainder. The horizontal dashed line denotes the 7.5% 10-year risk threshold recommended by ACC/AHA guidelines for consideration of statin therapy initiation.

**Figure 2.**
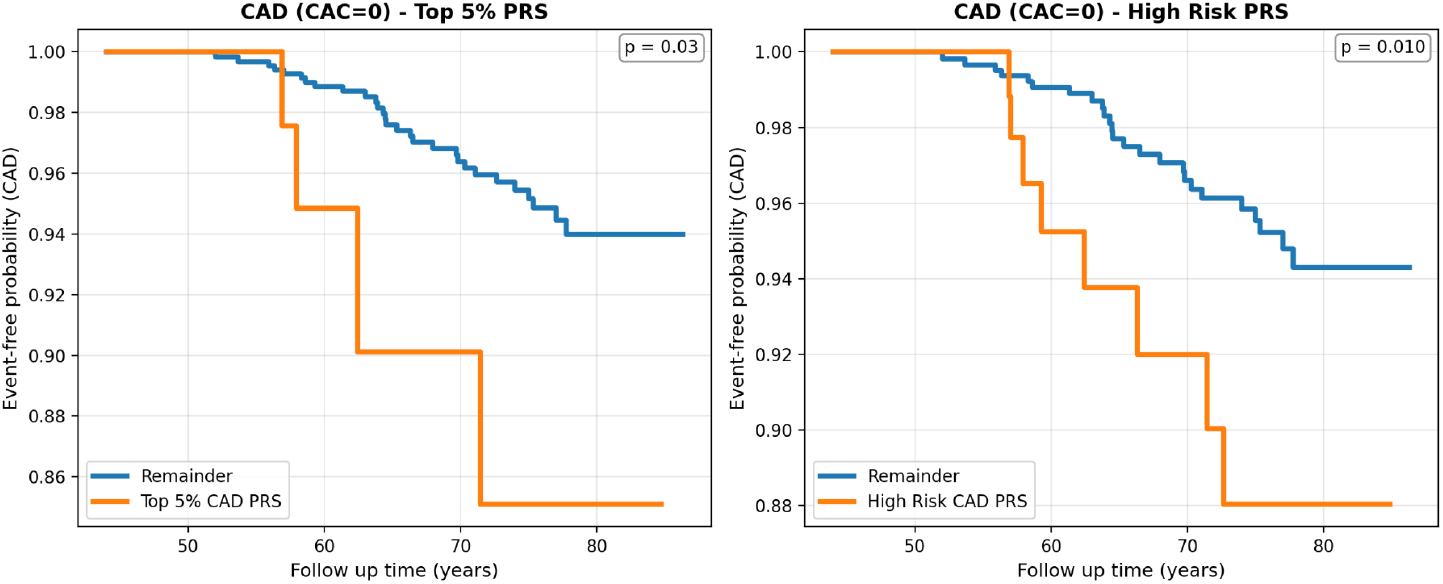
Kaplan–Meier estimates of CAD event-free survival during full follow-up among MESA participants with baseline CAC=0. Kaplan–Meier estimates of CAD event–free survival over the entire study follow-up among MESA participants with baseline CAC=0. Participants were stratified using two PRS-based definitions derived from the 8 Billion framework: (1) Top 5% PRS versus remainder of the cohort; and (2) High-Risk PRS versus remainder. The high-risk classification was defined at the individual level because 8 Billion calibrates PRS using a genetically similar reference neighborhood specific to each participant; therefore, a single universal threshold is not applicable across individuals. Curves depict cumulative event-free survival for incident CAD during the full follow-up period.

**Table 1.** Distribution of cases and controls across ancestry groups. The table summarizes the number of incident CAD cases and controls within each ancestry category. Due to the very low number of events observed in East Asian (EAS) and Admixed American (AMR) participants after application of inclusion criteria, these groups were excluded from the primary analyses to ensure statistical stability and reliability of estimates.

| Ancestry | Cases | Controls | Total |
| --- | --- | --- | --- |
| European | 17 | 792 | 809 |
| African | 5 | 407 | 412 |
| Admixed American | 2 | 278 | 280 |
| East Asian | 1 | 281 | 282 |
| Admixed | 11 | 468 | 479 |

**Table 2.**
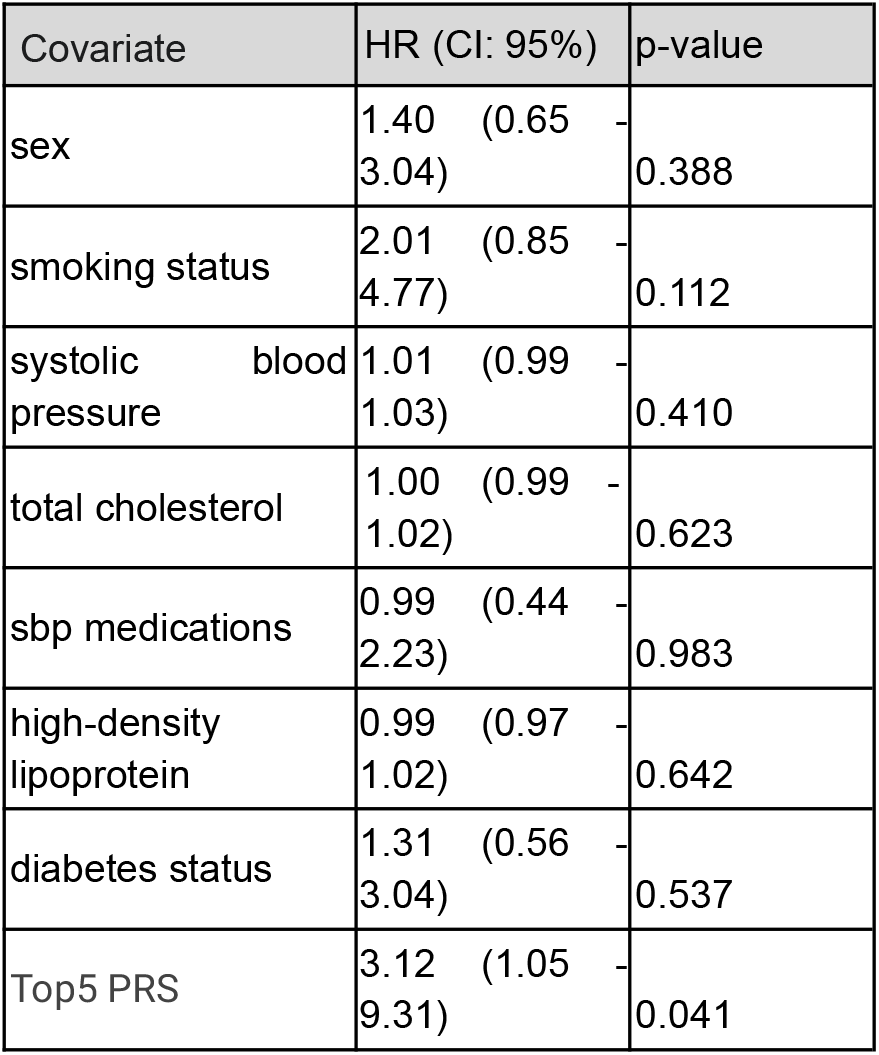
Association between Top 5% PRS and incident coronary artery disease (CAD) among MESA participants with baseline CAC=0. Hazard ratios (HRs) were estimated using Cox proportional hazards models to evaluate the association between classification in the Top 5% PRS group and incident CAD events. The Top 5% PRS group was defined as individuals whose PRS-derived odds ratios ranked in the highest 5% of the overall cohort. Models were adjusted for principal components of ancestry (PC1–PC4), sex, smoking status, systolic blood pressure, total cholesterol, HDL cholesterol, diabetes, and antihypertensive medication use. Continuous covariates were standardized within ancestry groups. Hazard ratios, 95% confidence intervals, and corresponding P values are presented for each covariate.

| Covariate | HR (CI: 95%) | p-value |
| --- | --- | --- |
| sex | 1.40 (0.65 - 3.04) | 0.388 |
| smoking status | 2.01 (0.85 - 4.77) | 0.112 |
| systolic blood pressure | 1.01 (0.99 - 1.03) | 0.410 |
| total cholesterol | 1.00 (0.99 - 1.02) | 0.623 |
| sbp medications | 0.99 (0.44 - 2.23) | 0.983 |
| high-density lipoprotein | 0.99 (0.97 - 1.02) | 0.642 |
| diabetes status | 1.31 (0.56 - 3.04) | 0.537 |
| Top5 PRS | 3.12 (1.05 - 9.31) | 0.041 |

**Table 3.** Association between high polygenic risk and incident coronary artery disease (CAD) among MESA participants with baseline CAC=0. Hazard ratios (HRs) were estimated using Cox proportional hazards models to evaluate the association between classification in the High-Risk PRS group and incident CAD events. The High-Risk PRS threshold was defined at the individual level using the 8 Billion framework, which calibrates PRS based on a genetically similar reference neighborhood for each participant; therefore, a single fixed threshold was not applied across the cohort. Models were adjusted for principal components of ancestry (PC1–PC4), sex, smoking status, systolic blood pressure, total cholesterol, HDL cholesterol, diabetes, and antihypertensive medication use. Continuous covariates were standardized within ancestry groups. Hazard ratios, 95% confidence intervals, and corresponding P values are shown for each covariate.

| Covariate | HR (CI: 95%) | p-value |
| --- | --- | --- |
| sex | 1.40 (0.65 - 3.04) | 0.389 |
| smoking status | 1.97 (0.83 - 4.65) | 0.124 |
| systolic blood pressure | 1.01 (0.99 - 1.03) | 0.440 |
| total cholesterol | 1.00 (0.99 - 1.01) | 0.695 |
| sbp medications | 1.01 (0.45 - 2.28) | 0.985 |
| high-density lipoprotein | 0.99 (0.97 - 1.02) | 0.618 |
| diabetes status | 1.27 (0.54 - 2.96) | 0.580 |
| High Risk PRS | 2.52 (1.12 - 5.66) | 0.025 |

**Table 4.** Association between Top 5% PRS and incident CAC ≥100 among MESA participants with baseline CAC=0. Hazard ratios (HRs) were estimated using Cox proportional hazards models to evaluate the association between classification in the Top 5% PRS group and development of significant coronary calcification (CAC ≥100). The Top 5% PRS group was defined as individuals whose PRS-derived odds ratios ranked in the highest 5% of the overall cohort. Models were adjusted for principal components of ancestry (PC1–PC4), sex, smoking status, systolic blood pressure, total cholesterol, HDL cholesterol, diabetes, and antihypertensive medication use. Continuous covariates were standardized within ancestry groups. Hazard ratios, 95% confidence intervals, and corresponding P values are shown for each covariate.

| Covariate | HR (CI: 95%) | p-value |
| --- | --- | --- |
| sex | 1.41 (0.87 - 2.30) | 0.163 |
| smoking status | 2.47 (1.46 - 4.18) | 0.001 |
| systolic blood pressure | 1.01 (1.00 - 1.02) | 0.240 |
| total cholesterol | 1.00 (0.99 - 1.01) | 0.684 |
| sbp medications | 2.03 (1.28 - 3.24) | 0.003 |
| high-density lipoprotein | 0.99 (0.97 - 1.01) | 0.287 |
| diabetes status | 2.01 (1.24 - 3.26) | 0.005 |
| Top5 PRS | 1.50 (0.60 - 3.75) | 0.386 |

**Table 5.** Association between high polygenic risk and incident CAC ≥100 among MESA participants with baseline CAC=0. Hazard ratios (HRs) were estimated using Cox proportional hazards models to evaluate the association between classification in the High-Risk PRS group and development of significant coronary calcification (CAC ≥100). The High-Risk PRS threshold was defined at the individual level using the 8 Billion framework, which calibrates PRS based on a genetically similar reference neighborhood for each participant; therefore, a single fixed threshold was not applied across the cohort. Models were adjusted for principal components of ancestry (PC1–PC4), sex, smoking status, systolic blood pressure, total cholesterol, HDL cholesterol, diabetes, and antihypertensive medication use. Continuous covariates were standardized within ancestry groups. Hazard ratios, 95% confidence intervals, and corresponding P values are shown for each covariate.

| Covariate | HR (CI: 95%) | p-value |
| --- | --- | --- |
| sex | 1.41 (0.87 - 2.30) | 0.164 |
| smoking status | 2.45 (1.45 - 4.15) | 0.001 |
| systolic blood pressure | 1.01 (1.00 - 1.02) | 0.255 |
| total cholesterol | 1.00 (0.99 - 1.01) | 0.702 |
| sbp medications | 2.05 (1.28 - 3.27) | 0.003 |
| high-density lipoprotein | 0.99 (0.97 - 1.01) | 0.275 |
| diabetes status | 1.99 (1.23 - 3.23) | 0.005 |
| High Risk PRS | 1.18 (0.62 - 2.23) | 0.610 |

When CAC progression to ≥100 was evaluated, associations with PRS were attenuated and not statistically significant after adjustment (Top 5%: HR, 1.50; 95% CI, 0.60–3.75; P=0.386; high-risk PRS: HR, 1.18; 95% CI, 0.62–2.23; P=0.61). In contrast, smoking, antihypertensive therapy, and diabetes were independently associated with CAC progression.

## Discussion

In this multi-ethnic cohort of individuals with baseline CAC=0, the Allelica Multi-ancestry CAD PRS, calculated using the 8 Billion framework, stratified risk of incident coronary events independently of traditional cardiovascular risk factors. Individuals in the highest PRS strata had higher rates of clinical CAD despite identical baseline imaging findings. These data inform the clinical interpretation of a CAC score of 0. Although CAC=0 is generally associated with low short-to intermediate-term risk, risk was heterogeneous within this group. Elevated polygenic burden identified a subset with higher subsequent event rates, whereas individuals with lower PRS had lower observed risk. Incorporation of genetic risk may therefore assist in distinguishing patients with persistently low risk from those with residual risk not captured by calcified plaque assessment alone. In fully adjusted models, PRS remained independently associated with incident CAD, whereas conventional risk factors were not statistically significant within this CAC=0 subset. This suggests that polygenic risk captures an additional dimension of atherosclerotic susceptibility not reflected by standard clinical variables or calcified plaque burden. Notably, PRS was not significantly associated with progression to CAC ≥100 after multivariable adjustment. This apparent dissociation between clinical events and calcification progression is biologically plausible. CAC reflects calcified plaque burden, whereas early and potentially vulnerable atherosclerotic lesions are frequently noncalcified. Multiple imaging studies have demonstrated that individuals with high polygenic risk exhibit a greater burden and faster progression of noncalcified, lipid-rich plaques features associated with plaque vulnerability and acute coronary syndromes[22,23,24]. Accordingly, PRS may preferentially identify susceptibility to high-risk, noncalcified plaque rather than to calcification per se. In individuals with CAC=0 and elevated PRS, coronary computed tomographic angiography (CCTA) could therefore be considered to evaluate noncalcified plaque burden when clinically appropriate. By applying the 8 Billion framework, which models genetic similarity as a continuum rather than assigning individuals to rigid ancestry categories, we successfully stratified risk in this multi-ethnic cohort. This approach enabled individualized PRS calibration across African, European, and admixed participants and supports the feasibility of genetically informed risk assessment in heterogeneous real-world populations.

## Conclusions

Genetic risk assessment improved cardiovascular risk stratification among individuals with CAC=0. High CAD PRS identified a subgroup at substantially elevated risk of incident CAD despite absence of baseline calcification, whereas non elevated PRS reinforced the protective implication of CAC=0. These findings support consideration of elevated polygenic risk as a risk-enhancing factor in preventive decision-making, suggest a potential role for adjunctive imaging such as CCTA in selected high-PRS individuals with CAC=0, and provide a biologically plausible explanation, rooted in inherited susceptibility, for residual events observed in patients with a zero CAC score.

## Data Availability

Data available through the MESA public repository

https://mesa-nhlbi.org/

## Non-standard Abbreviations and Acronyms

ACC: American College of Cardiology AHA American Heart Association AMR Admixed American
ASCVD: atherosclerotic cardiovascular disease CAC coronary artery calcium
CAD: coronary artery disease CI confidence interval
CVD: cardiovascular disease EAS East Asian
HDL: high-density lipoprotein HR hazard ratio
MESA: Multi-Ethnic Study of Atherosclerosis OR odds ratio
PC: principal component
PCE: Pooled Cohort Equations PRS polygenic risk score
SBP: systolic blood pressure

## References

1. Lozano R, Naghavi M, Foreman K, et al. Global and regional mortality from 235 causes of death for 20 age groups in 1990 and 2010: a systematic analysis for the Global Burden of Disease Study 2010 Lancet. 2012; 380: 2095–2128.

2. Khera AV, Emdin CA, Drake I, Natarajan P, Bick AG, Cook NR, et al. Genetic Risk, Adherence to a Healthy Lifestyle, and Coronary Disease. N Engl J Med. 2016; 375: 2349–2358.

3. Said MA, Verweij N, van der Harst P. Associations of Combined Genetic and Lifestyle Risks With Incident Cardiovascular Disease and Diabetes in the UK Biobank Study. JAMA Cardiol. 2018; 3: 693–702.

4. SCORE2 working group and ESC Cardiovascular risk collaboration. SCORE2 risk prediction algorithms: new models to estimate 10-year risk of cardiovascular disease in Europe. Eur Heart J. 2021; 42: 2439–2454.

5. Wilson PW, D’Agostino RB, Levy D, Belanger AM, Silbershatz H, Kannel WB. Prediction of coronary heart disease using risk factor categories. Circulation. 1998; 97: 1837–1847.

6. Goff DC Jr, Lloyd-Jones DM, Bennett G, Coady S, D’Agostino RB, Gibbons R, et al. American College of Cardiology/American Heart Association Task Force on Practice Guidelines. 2013 ACC/AHA guideline on the assessment of cardiovascular risk: a report of the American College of Cardiology/American Heart Association Task Force on Practice Guidelines. Circulation. 2014; 129: S49–73.

7. Michos ED, Nasir K, Braunstein JB, Rumberger JA, Budoff MJ, Post WS, et al. Framingham risk equation underestimates subclinical atherosclerosis risk in asymptomatic women. Atherosclerosis. 2006; 184: 201–206.

8. Agatston AS, Janowitz WR, Hildner FJ, Zusmer NR, Viamonte M Jr, Detrano R. Quantification of coronary artery calcium using ultrafast computed tomography. J Am Coll Cardiol. 1990; 15: 827–832.

9. Detrano R, Guerci AD, Carr JJ, et al. Coronary calcium as a predictor of coronary events in four racial or ethnic groups. N Engl J Med. 2008; 358: 1336–1345.

10. Polonsky TS, McClelland RL, Jorgensen NW, et al. Coronary artery calcium score and risk classification for coronary heart disease prediction. JAMA. 2010; 303: 1610–1616.

11. Erbel R, Möhlenkamp S, Moebus S, et al. Coronary risk stratification, discrimination, and reclassification improvement based on quantification of subclinical coronary atherosclerosis: the Heinz Nixdorf Recall study. J Am Coll Cardiol. 2010; 56: 1397–1406

12. Raggi P, Shaw LJ, Berman DS, Callister TQ. Prognostic value of coronary artery calcium screening in subjects with and without diabetes. J Am Coll Cardiol. 2004; 43: 1663–1669.

13. Paović J, Bos D, Ikram MK, Ikram MA, Kavousi M, Leening MJG. Guideline-Directed Application of Coronary Artery Calcium Scores for Primary Prevention of Atherosclerotic Cardiovascular Disease. JACC Cardiovasc Imaging. 2025; 18: 465–475.

14. Maron DJ, Budoff MJ, Sky JC, Bommer WJ, Epstein SD, Fisher DA, Stock EO, Taylor AJ, Wong ND, DeMaria AN. Coronary Artery Calcium Staging to Guide Preventive Interventions: A Proposal and Call to Action. JACC Adv. 2024; 3: 101287.

15. Nasir K, Cainzos-Achirica M. Role of coronary artery calcium score in the primary prevention of cardiovascular disease. BMJ. 2021; 373:n776.

16. Liew G, Chow C, van Pelt N, et al. Cardiac Society of Australia and New Zealand Position Statement: Coronary Artery Calcium Scoring. Heart Lung Circ. 2017; 26: 1239–1251.

17. Sama C, Abdelhaleem A, Velu D, et al. Non-calcified plaque in asymptomatic patients with zero coronary artery calcium score: A systematic review and meta-analysis. J Cardiovasc Comput Tomogr. 2024; 18: 43–49.

18. Busby GB, Kulm S, Bolli A, Kintzle J, Domenico PD, Bottà G. Ancestry-specific polygenic risk scores are risk enhancers for clinical cardiovascular disease assessments. Nat Commun. 2023; 14: 7105.

19. The data/analyses presented in the current publication are based on the use of study data downloaded from the dbGaP web site, under https://dbgap.ncbi.nlm.nih.gov/beta/study/phs000420.v6.p3/#study

20. McPherson R, Tybjaerg-Hansen A. Genetics of Coronary Artery Disease. Circ Res. 2016; 118: 564–578.

21. Wong CJ, Inouye L. What’s in a Number? Risk Thresholds in Different Statin Guidelines. J Gen Intern Med. 2017 Oct;32(10):1071–1073. doi: 10.1007/s11606-017-4114-y. Epub 2017 Jun 29. PMID: 28664259; PMCID: PMC5602765.

21. van Veelen, A.; van der Sangen, N.M.Rx.; Delewi, R.; Beijk, M.A.M.; Henriques, J.P.S.; Claessen, B.E.P.M. Detection of Vulnerable Coronary Plaques Using Invasive and Non-Invasive Imaging Modalities. J. Clin. Med. 2022, 11, 1361. 10.3390/jcm11051361

22. Cornelissen A, Gadhoke NV, Ryan K, Hodonsky CJ, Mitchell R, Bihlmeyer NA, Duong T, Chen Z, Dikongue A, Sakamoto A, Sato Y. Polygenic risk score associates with atherosclerotic plaque characteristics at autopsy. Arteriosclerosis, thrombosis, and vascular biology. 2024 Jan;44(1):300–13

23. Nurmohamed NS, Shim I, Gaillard EL, Ibrahim S, Bom MJ, Earls JP, Min JK, Planken RN, Choi AD, Natarajan P, Stroes ES. Polygenic risk is associated with long-term coronary plaque progression and high-risk plaque. Cardiovascular Imaging. 2024 Dec 1;17(12):1445–59.

24. Petranovic M, Lai YP, Huck D, Shiyovich A, Besser S, Freire CV, Berman A, Weber B, Fahed A, Miao J, Hainer J. POLYGENIC RISK SCORE FOR CORONARY ARTERY DISEASE PREDICTS PRESENCE, BURDEN, AND SEVERITY OF ATHEROSCLEROTIC PLAQUE IN PATIENTS UNDERGOING CORONARY CT ANGIOGRAPHY. American Journal of Preventive Cardiology. 2025 Sep 1;23:101212.

